# Time-series clustering for home dwell time during COVID-19: what can we learn from it?

**DOI:** 10.1101/2020.09.27.20202671

**Authors:** Xiao Huang, Zhenlong Li, Junyu Lu, Sicheng Wang, Hanxue Wei, Baixu Chen

## Abstract

In this study, we investigate the potential driving factors that lead to the disparity in the time-series of home dwell time, aiming to provide fundamental knowledge that benefits policy-making for better mitigation strategies of future pandemics. Taking Metro Atlanta as a study case, we perform a trend-driven analysis by conducting Kmeans time-series clustering using fine-grained home dwell time records from SafeGraph, and further assess the statistical significance of sixteen demographic/socioeconomic variables from five major categories. We find that demographic/socioeconomic variables can explain the disparity in home dwell time in response to the stay-at-home order, which potentially leads to disparate exposures to the risk from the COVID-19. The results further suggest that socially disadvantaged groups are less likely to follow the order to stay at home, pointing out the extensive gaps in the effectiveness of social distancing measures exist between socially disadvantaged groups and others. Our study reveals that the long-standing inequity issue in the U.S. stands in the way of the effective implementation of social distancing measures. Policymakers need to carefully evaluate the inevitable trade-off among different groups, making sure the outcomes of their policies reflect interests of the socially disadvantaged groups.

**Highlights:**

- We perform a trend-driven analysis by conducting Kmeans time-series clustering using fine-grained home dwell time records from SafeGraph.
- We find that demographic/socioeconomic variables can explain the disparity in home dwell time in response to the stay-at-home order.
- The results suggest that socially disadvantaged groups are less likely to follow the order to stay at home, potentially leading to more exposures to the COVID-19.
- Policymakers need to make sure the outcomes of their policies reflect the interests of the disadvantaged groups.

## Introduction

The coronavirus disease 2019 (COVID-19) is a global threat that raises worldwide concerns with escalating economic, social, and health challenges. On March 11, the World Health Organization (WHO) officially declared COVID-19 as a pandemic, pointing to the sustained risk of further global spread and urging countries and regions to join forces [1]. As of September 6 (the time of writing), there had been a total of 26,763,217 infections and 876,616 deaths globally, and the U.S. accounts for 23.0% of the global infections and 21.3% of the global deaths [2]. We are still witnessing widespread community transmission of the COIVD-19 all over the world. Unfortunately, to date, there is neither a vaccine nor a pharmacological agent found to reduce the transmission of severe acute respiratory syndrome coronavirus-2 (SARS-CoV-2), the virus that causes COVID-19 [3].

In response to the threat of COVID-19, social distancing measures are one of the primary tools to reduce the transmission of the SARS-CoV-2 virus. National and local governments have promoted stay-at-home orders while required non-essential business closures to reduce the risk of transmission by further enhancing social distancing measures [4]. Studies have found that such early governmental policies have been proved rather effective in China [5], Korea[6], and many European countries [7,8], as notable declines in transmission rates were observed following the implementation of strong mobility-reducing measures. In the U.S., many states, counties, and cities began issuing stay-at-home or similar mitigation measures that require residents to reduce movement and stay home as early as in March 2020, which leads to a considerable increase in home dwell time. The earliest stay-at-home order was implemented in the Bay Area, CA, on March 16, 2020, and soon after (3 days later), a state-wide stay-at-home order was issued in CA [9]. Gradually, an increasing number of states started to adopt this strategy. By March 24, more than 50% of the U.S. population was under a stay-at-home order, and this number soared from 50% to 95% by April 4 [9]. Despite the widely-adopted stay-at-home orders, there was mounting evidence of the disparate responses that potentially leaves vulnerable populations unequally exposed to the COVID-19 pandemic [10,11]. When facing the same mitigation measures, such disparate responses from residents are largely due to the different socioeconomic status, reflecting the long-standing problem of health inequity in the U.S., which usually exaggerates the consequences from disproportionate responses by inflicting long-term negative outcomes for the socially-disadvantaged groups [12]. Thus, identifying demographic and socioeconomic variables that potentially drive the disparity in the implementation of stay-at-home orders deserves much attention.

Many studies have investigated the disparity in response to the stay-at-home orders during the COVID-19 pandemic. The responses are usually quantified by home dwell time, travel distance, and POI (point of interest) visits, thanks to the availability of mobility datasets that facilitate the rapid monitoring of human mobility. Chiou and Tucker [13] investigated the U.S. tract-level correlation between income and self-isolation at home and found that high-income earners generally spend more time at home (their evidence points out that the access to high-speed Internet plays an important role). Barnett-Howell and Mobarak [14] found that people with less income tend to place greater value on their livelihood concerns than contracting COVID-19, consequently resulting in smaller epidemiological and economic benefits of social distancing measures in poorer regions. Jones [15] documented the urban-rural discrepancy in threat awareness, as 54% of urban residents in the states view the COVID-19 as a major threat, compared with 42% of those living in the suburbs and just 27% of rural residents in the same states. This urban-rural disparity in risk awareness presumably drives the disparity in their daily movement, therefore further translates to the disparity in COVID-19 exposure. Via the investigation of U.S. county-level mobility records, Lou et al. [16] found differential impacts of stay-at-home orders on economics groups, where the lower-income group is less likely to follow the order to stay at home, evidenced by their longer travel distances compared with the higher-income group. Similarly, Huang et al. [10] compared four popular mobility datasets and found that, regardless of their unique characteristics, all selected mobility datasets suggest a statistically significant positive correlation between mobility reduction and income at the U.S. county scale. Despite the above efforts, the soundness of correlating disparity in response to demographic/socioeconomic variables is hampered by the coarse geographical units, as mitigation policies may vary in different countries, states, and even counties; therefore, the documented disparity in response may result from the discrepancy in mitigation policies, not from the varying demographic/socioeconomic indicators. Thus, the examination of fine-grained mobility records (e.g., at the census tract or block group level) are in great need. In addition, most existing studies utilize indices summarized during a specific period to quantify the mobility-related response, neglecting the dynamic perspectives revealed from time-series data. In comparison, time-series trend-based analytics may provide valuable insights in distinguishing different dynamic patterns of mobility records, thus warranting further investigation.

The objective of this study is to explore the capability of time-series clustering in categorizing fine-grained mobility records during the COVID-19 pandemic, and further investigate what demographic/socioeconomic variables differ among the categories with statistical significance. Taking advantage of the home dwell time at Census Block Group (CBG) level from the SafeGraph [17], and using the Atlanta-Sandy Springs-Roswell metropolitan statistical area (MSA) (hereafter referred to as Metro Atlanta) as a study case, this study investigates the potential driving factors that lead to the disparity in the time-series of home dwell time during the COVID-19 pandemic, providing fundamental knowledge that benefits policy-making for better mitigation measures of future pandemics. The contributions of this work are summarized as follows:

- We perform a trend-driven analysis by conducting Kmeans time-series clustering using fine-grained home dwell time records from SafeGraph.
- We assess the statistical significance of sixteen selected demographic/socioeconomic variables among categorized groups derived from the time-series clustering. Those variables cover economic status, races and ethnicities, age and household type, education, and transportation.
- We discuss the potential demographic/socioeconomic variables that lead to the disparity in home dwell time during the COVID-19 pandemic, how they reflect the long-standing health inequity in the U.S., and what can be suggested for better policy-making.

The remainder of the paper is organized as follows. Section 2 introduces the datasets used in this study. Section 3 presents the methodological approaches we applied. Section 4 describes the contexts of the study case (Metro Atlanta). Section 5 presents the results of time-series clustering, the results of the analysis of variance, and the discussion. Section 6 concludes our article.

## 2. Datasets

### 3.1. Home dwell time

The home dwell time records are derived from SafeGraph (https://www.safegraph.com/), a data company that aggregates anonymized location data from numerous applications in order to provide insights about physical places. SafeGraph aggregates data using a panel of GPS points from anonymous mobile devices and determines the home location as the common nighttime location of each mobile device over a six-week period to a Geohash-7 granularity (∼153m × ∼ 153m) [17]. To enhance privacy, SafeGraph excludes CBG information if fewer than five devices visited an establishment in a month from a given CBG. The data records used in this study are the median home dwell time in minutes for all devices with a certain CBG on a daily basis. For each device, the observed minutes at home across the day are summed, and the median value for all devices with a certain CBG is further calculated [17]. The raw SafeGraph dataset we used for the year 2020 spans from January 1, 2020, to August 31, 2020 (244 days) with daily home dwell records (in mins) for a total of 219,972 CBGs. Heat map of home dwell time for these CBGs are presented in Figure 1. The impact of COVID-19 can be observed, as home dwell time notably increased after the declaration of National Emergency on March 13, 2020 [18] (Figure 1), despite the disparity in the increasing intensity. After the lifting of strict social distancing measures in early May, however, home dwell time starts to decrease and returns to the pre-pandemic level (Figure 1). The increased variation of home dwell time after the National Emergency declaration indicates that CBGs have different responses to the pandemic and the government order. Despite the large number of CBGs, not all CBGs contain sufficient records to derive stable time-series that can be used for clustering. The details of the preprocessing steps are presented in Section 3.1.

**Figure 1.**
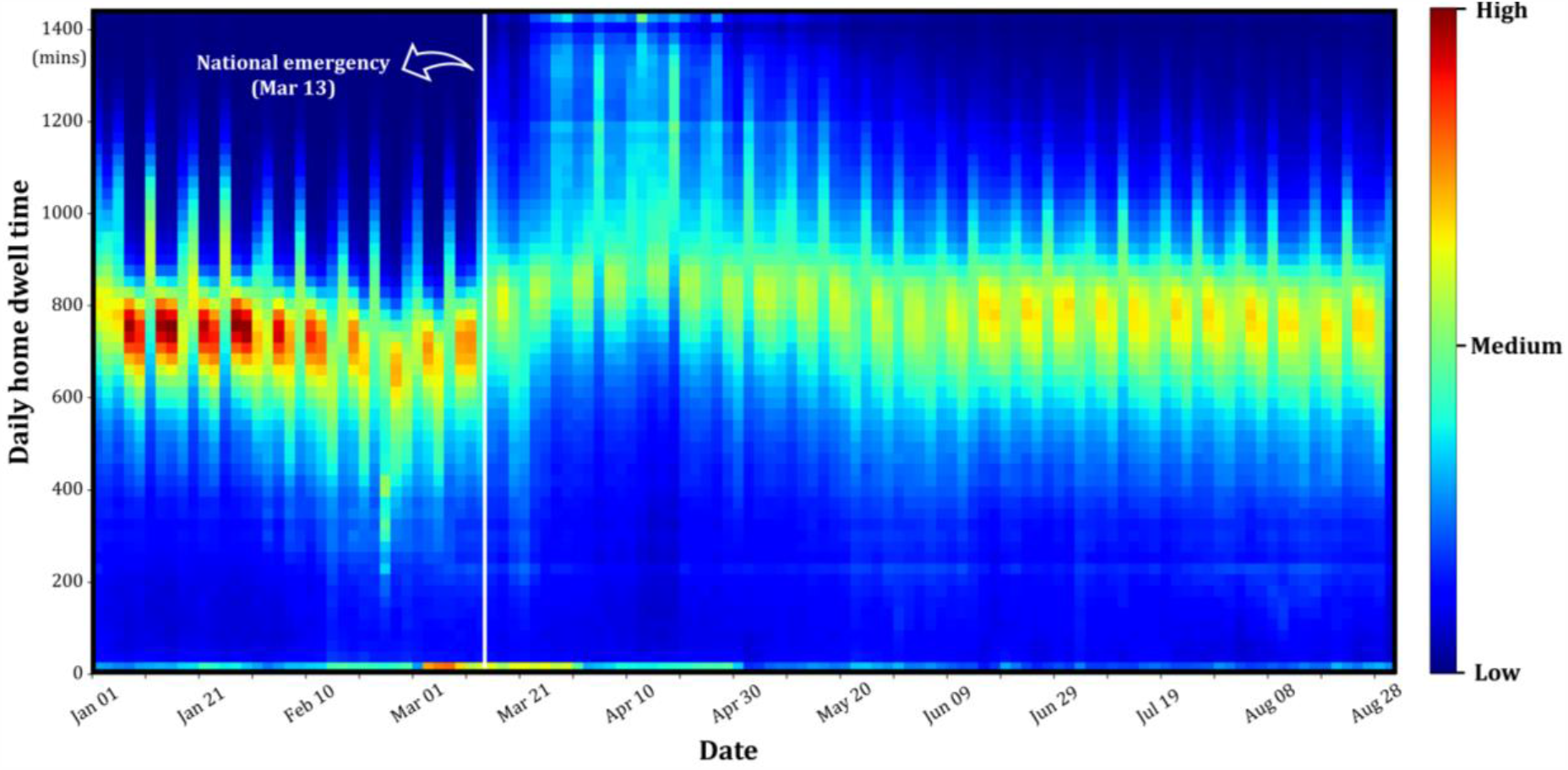
Heat map of home dwell time for 219,972 CBGs in the U.S. from January 1, 2020, to August 31, 2020. High/low concentrations are marked as red/blue.

### 3.2. Demographic/socioeconomic variables

Demographic and socioeconomic variables in this study are derived from the American Community Survey (ACS), collected by the U.S. Census Bureau. ACS is an ongoing nationwide survey that investigates a variety of aggregated information about U.S. residents at different geographic levels every year [19]. ACS randomly selects monthly samples based on housing unit addresses and publishes annual estimates datasets (i.e., 12-month samples). In addition to the 1- year datasets, ACS also releases 3-year estimates (i.e., 36-month samples) and 5-year estimates (i.e., 60-month samples). Compared to the 1-year and 3-year datasets, 5-year estimates cover the most areas, have the largest sample size, and contain the most reliable information [20]. In this study, we use the latest 5-year ACS data, i.e., the 2014-2018 ACS 5-year estimates, obtained from Social Explorer (https://www.socialexplorer.com/). We recode the variables from ACS data as five major categories: 1) Economic status; 2) Races and ethnicities; 3) Gender, age and household type; 4) Education; 5) Transportation. Previous empirical studies suggested that these variables could be associated with the pattern of daily travels and participation of out-of-home activities [21–24]. The detailed information of the variables within the five categories is presented in Table 1. In addition, CBG boundaries are derived from 2019 TIGER/Line Shapefiles by U.S. Census Bureau (https://www.census.gov/cgi-bin/geo/shapefiles/index.php).

**Table 1.** Notations and descriptions of the demographic and socioeconomic variable in the selected five major categories.

| Variable notations | Descriptions |
| --- | --- |
| **Economic status** |  |
| pct_low_income | percent of household income less than \$15,000 |
| pct_high_income | percent of household income greater than \$150,000 |
| median_hhinc | median household income |
| pct_unemployrate | unemployment rate |
| **Races and ethnicities** |  |
| pct_white | percent of White |
| pct_black | percent of Black |
| pct_hispanic | percent of Hispanic |
| **Gender, age and household type** |  |
| pct_female | percent of female |
| pct_elderly | percent of age 65 or older |
| pct_signleparent | percent of single-parent families among parenting families having children under 18 |
| **Education** |  |
| pct_low_edu | percent of education equal or less than highschool |
| pct_grad_edu | percent of education of master, professional, or doctoral degrees |
| **Transportation** |  |
| pct_workhome | percent of work from home |
| pct_short_commute | percent of commuters with 10 min or shorter trips |
| pct_long_commute | percent of commuters with 40 min or longer trips |
| pct_0car | percent of 0 car households |

## 3. Methods

### 3.1. Preprocessing

Several preprocessing steps are applied to ensure that CBGs within the study area contain sufficient and valid records to derive stable time-series that can be used for clustering. We first select CBGs that fall within the study area, i.e., Metro-Atlanta (more details of the Metro-Atlanta can be found in Section 4), which results in a total of 2,687 CBGs. As SafeGraph uses digital devices to measure home dwell time, the number of available devices in each CBG greatly determines the representativeness and the stability of the time-series. We plot the spatial distribution of median daily device count within the Metro Atlanta area and observe that CBGs dominated by non-residential zones tend to have less daily device count (Figure 2a), presumably due to the low number of home locations identified via SafeGraph’s algorithm (see Section 3.1). We keep CBGs with more than 200 days (out of 244 days) of home dwell time records to ensure reliable time-series can be generated. To fill the missing data, we adopt the approach from Huang et al. [10], where missing data are filled via a simple linear interpolation by assuming that home dwell time changes linearly between two consecutive available records. Our preliminary investigation suggests that stable time-series of daily home dwell time can be achieved when daily device count reaches 100. Thus, we calculate the median of daily device count for each CBG during the 244-day period and select CBGs with the median equal or larger than 100. We also observe that some CBGs present abnormal home dwell patterns with consecutive 0 values for a certain period of time. To avoid the potential problems caused by these CBGs on the performance of the clustering algorithm, we remove CBGs with 0 values that span more than three consecutive days. A total of 1,483 CBGs remain after the aforementioned preprocessing steps, and their representativeness is presented in Figure 2b. The representativeness is defined as the ratio between the median daily device count and the population from the ACS 2014-2018 estimates. The representativeness for most CBGs ranges from 5% - 10% (Figure 2b), which is considerably higher than Twitter [25], a commonly used open-sourced platform to derive mobility-related statistics.

**Figure 2.**
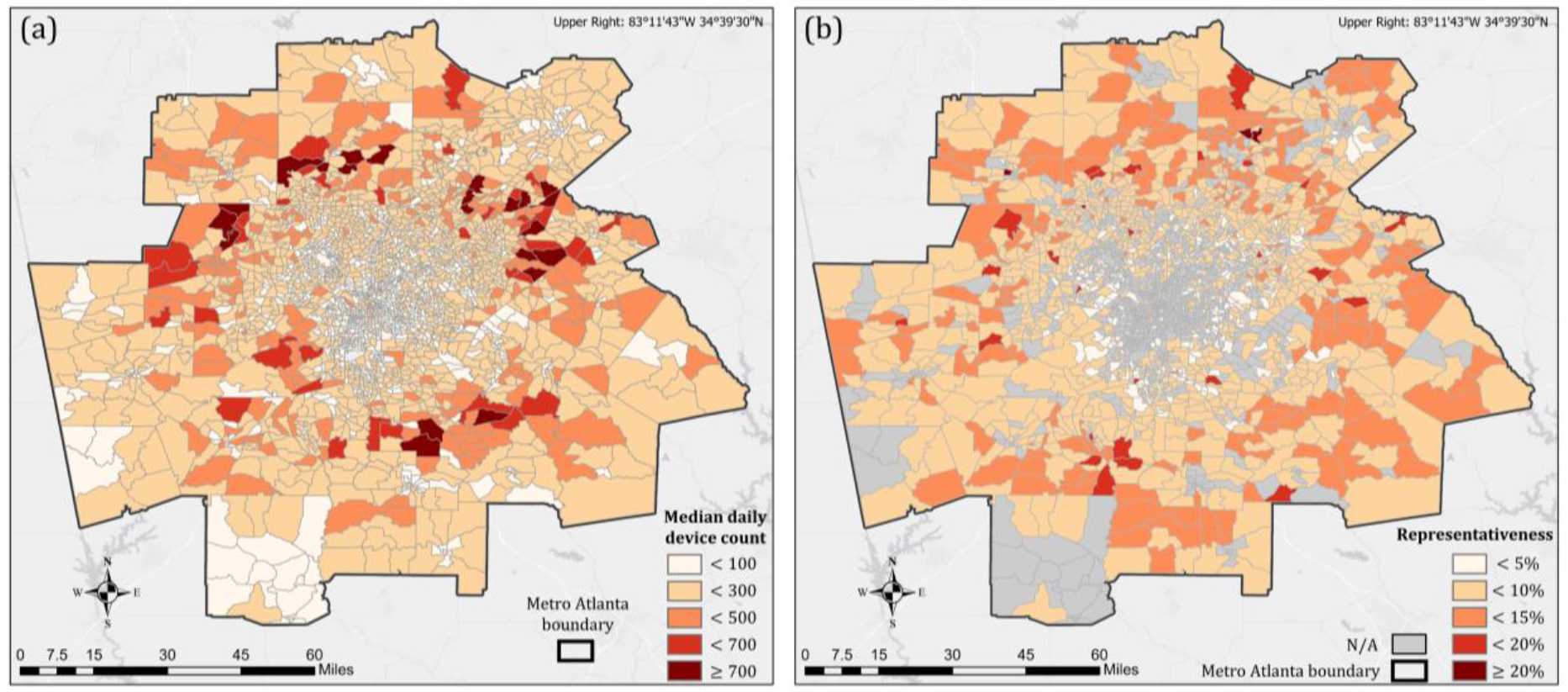
(a) The median daily device count from January 1 to August 31 at CBG level in Metro Atlanta; (b) Representativeness of each CBG in Metro Atlanta. The representativeness is defined as the ratio of the median daily device count to the population from the ACS 2014-2018 estimates. CBGs annotated with “N/A” fail to meet the requirements in the preprocessing steps and therefore are removed. CBG boundaries are derived from 2019 TIGER/Line Shapefiles.

### 3.2. Time-series clustering

Time-series clustering is the process of the partitioning a time-series dataset into a certain number of clusters, according to a certain similarity criterion. In this study, we aim to cluster the time-series of home dwell time in the CBGs within the study area. We adopt the design of K-means [26], an unsupervised partition-based clustering algorithm in which observations are categorized into the cluster with the nearest mean. The choice of similarity measurement in Kmeans is crucial to the detection of clusters [27]. Considering that the time-series of home dwell time for the majority of the CBGs present a similar shape but vary in intensity (Figure 1), we decide to calculate the Euclidean distance between two time-series.

Given a dataset on *n* time series *T* = {*t*_1_, *t*_1_, …, *t*_n_}, we aim to partition *T* into a total of *k* clusters, i.e., *C* = {*C*_1_, *C*_2_, … *C*_*k*_}, by minimizing the objective function J, given as:

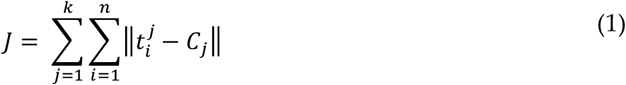

where *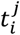* denotes the time-series *t* in category *j*, and ||·|| denotes the similarity measurement that measures the distance between *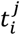* and the cluster center of *C*_*j*_ Let *t* and *C*_*j*_ each be a *m* - dimensional vector, where *m* equals the length of the time series (244 in this case). As Euclidean Distance is selected as similarity measurement in this study, 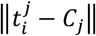 can be rewritten as:

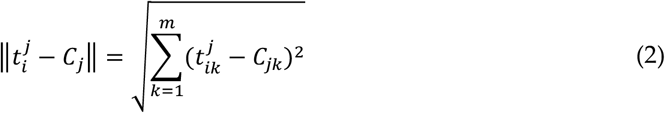

Further, Kmeans utilizes an iterative procedure with the following steps to derive the final category for each time-series candidate:

1. Initialize *k* cluster centroids *C*_1_, *C*_2_, … *C*_*k*_, arbitrarily.
2. Assign each time-series *t_i_* to its correct cluster *Cj*, according to *argmin* 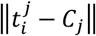
3. Update the centers *C*_*j*_ based on the new clusters.
4. Repeat steps 2 and 3 until convergence.

The Kmeans time-series clustering requires pre-specification of the total number of clusters (i.e., *k*), which inevitably introduces the subjective nature of deciding the constitution of reasonable clusters [28]. Through the investigation of the time-series dataset, we set *k* = 3, expecting to find three CBG clusters with different home dwell time patterns, following the stay-at-home order: 1) CBGs with a significant increase of home dwell time; 2) CBGs with a moderate increase of home dwell time; 3) CBGs with unnoticeable changes in home dwell time.

### 3.3. Analytical approaches

After the time-series clustering, three CBG clusters are therefore formed, each with a unique distribution pattern of daily home dwell time. Identifying the statistical difference in demographic/socioeconomic variables among these clusters facilitates a better understanding of what variables potentially lead to the disparity in home dwell time during the COVID-19 pandemic. Qualitatively, we label the CBG clusters, plot them spatially, and compare the spatial pattern of clusters with the spatial pattern of several major demographic/socioeconomic variables in the study area (see Figure 3 in Section 4). Quantitatively, we apply one-way ANOVA (Analysis of Variance) (α = 0.001) [29] to assess the statistical significance of five major indicators (see Table 1) among categorized CBG groups derived from the time-series clustering. As ANOVA does not provide insights into particular differences between pairs of cluster means, we further conduct Tukey’s test (α = 0.05, 0.01, 0.001) [30], a common and popular post-hoc analysis following ANOVA, to assess the statistical difference of demographic/socioeconomic variable between cluster pairs.

**Figure 3.**
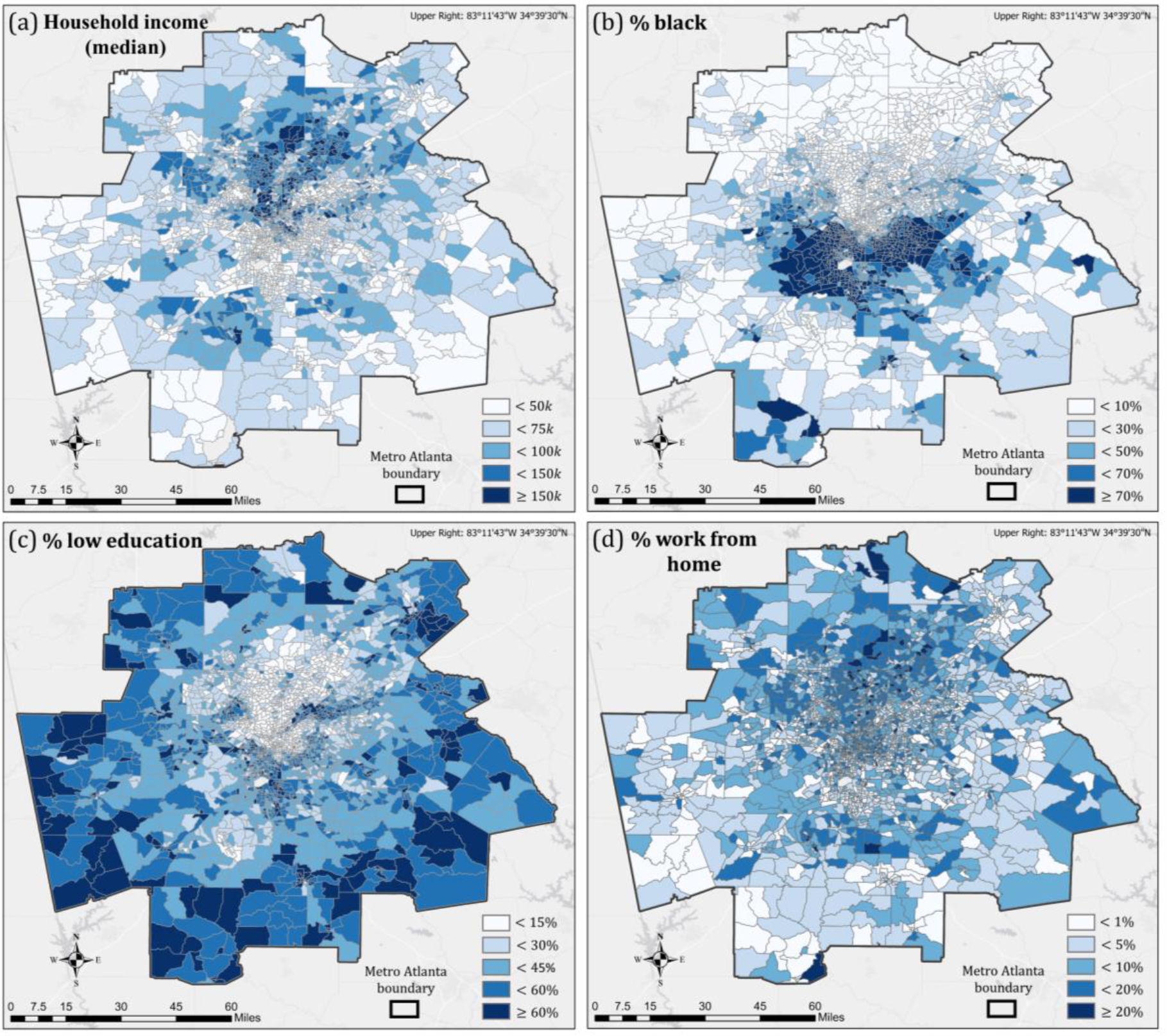
Profile of Metro Atlanta with four selected variables. (a) Median household income; (b) Percentage of black; (c) Percentage of low education (education equal or less than high school); (d) Percentage of workers who work from home. All statistics are derived from the ACS 2014- 2018 estimates. CBG boundaries are derived from 2019 TIGER/Line Shapefiles.

## 4. Profile of the study area

The study area defined in this study is referred to as Metro Atlanta, designated by the United States Office of Management and Budget (OMB) as the Atlanta–Sandy Springs–Alpharetta, Georgia (GA) Metropolitan Statistical Area (MSA). Metro Atlanta is the twelfth-largest MSA in the U.S. and the most populous metro area in GA [31]. The study area includes a total of 30 GA counties (listed in Table A) and has an estimated population of 5,975,424, according to the ACS 2014-2018 estimates. Metro Atlanta has grown rapidly since the 1940s. Despite its rapid growth, however, Metro Atlanta has shown widening disparities, including class and racial divisions, underlying the uneven growth and development, making it one of the metro regions with the most inequity [32–34]. It is the main reason why we chose this metro region to explore the disparity in responses to the COVID-19 pandemic. In the last few decades, the north metro area has absorbed most of the new growth, thanks to the northward shifting trend of the metro region’s white population and the rapid office, commercial, and retail development [35]. After the increasingly unbalanced development in recent decades, Metro Atlanta started to present a distinct north-south spatial disparity in many demographic/socioeconomic variables (Figure 3). Compared to the south metro region, the north region is characterized by higher income (Figure 3a), higher white percentages (Figure 3b), higher education (Figure 3c), and higher percentages of work-from-home workers (Figure 3d).

In contrast to the substantial spatial heterogeneity of socioeconomic status, GA’s governmental reactions to the COVID-19 pandemic are rather homogenous in space. On March 14, 2020, Governor Brian P. Kemp announced the public health state of emergency in GA. Twenty days later (April 3), the shelter-in-place order took effect for the entire state [36]. The strict social distancing measures lasted until late April when GA started to reopen gradually: resuming restaurant dine-in services (April 27), reopening bars and nightclubs with capacity limits (June 1), allowing the gatherings of 50 people (June 16), and reopening conventions and live performance (July 1) [37].

## 5. Results

### 5.1 Identified CBG clusters and their spatial distribution

Three CBG clusters are identified based on the time-series pattern in daily home dwell time via the Kmeans time-series clustering algorithm. CBGs in Cluster #1 are characterized by their unnoticeable changes in home dwell time throughout the entire time frame, suggesting that stay-at-home orders have a minimal effect on people living in these CBGs (Figure 4a). In comparison, CBGs in Cluster #2 (Figure 4b) and Cluster #3 (Figure 4c) responded to the strict measures implemented in March and April strongly. CBGs in Cluster #2 experienced a moderate increase in home dwell time during the implementation of strict social distancing measures (Figure 4b). Compared to Cluster #2 where the daily home dwell time increased up to 1,200 mins, CBGs in Cluster #3 saw a more dramatic increase, as the home dwell time for most of the CBGs in Cluster #3 reached 1,400 mins (out of 1440 mins in a day) in March and April, suggesting that mitigation measures have greatly changed people’s travel behavior in these CBGs (Figure 4c). Note that the three identified clusters are with different numbers of CBGs. Clusters #1, #2, and #3 have 157 CBGs, 778 CBGs, and 552 CBGs, respectively.

**Figure 4.**
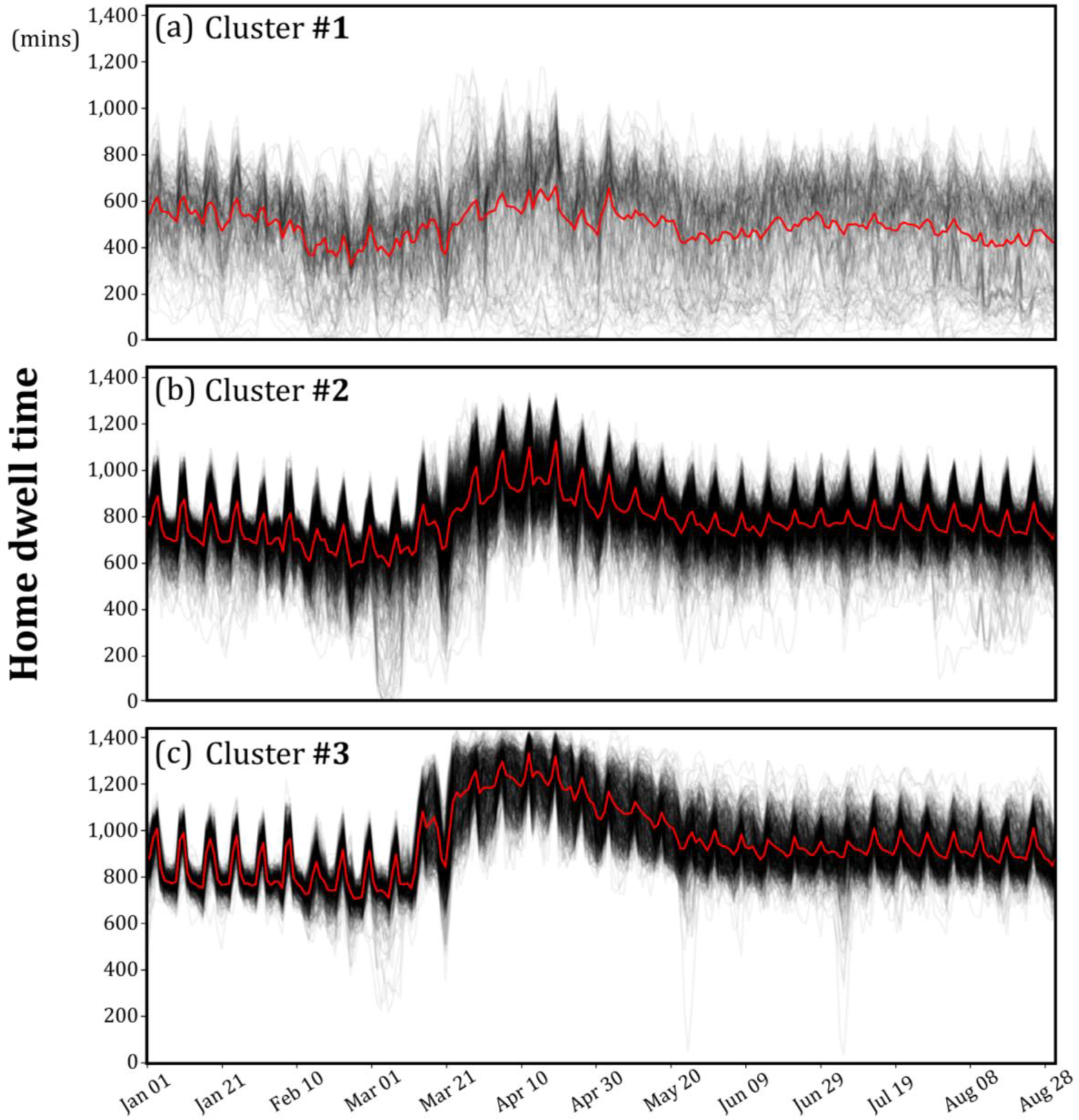
The time-series of the three identified CBG clusters. (a) Cluster #1: CBGs with unnoticeable changes in home dwell time (157 CBGs); (b) Cluster #2: CBGs with a moderate increase of home dwell time (778 CBGs); (c) Cluster #3: CBGs with a strong increase in home dwell time (552 CBGs).

Figure 5 shows the spatial distribution of the three CBG clusters, which presents a certain level of spatial autocorrelation, especially for Cluster #2 and Cluster #3. The Global Moran’s I [38] for the distribution of the three identified clusters is 0.243, and it is significant at the significance level of 0.01. In general, the spatial distribution implies that demographic/socioeconomic variables potentially drive the disparity in home dwell time during the pandemic. The distribution of CBGs in Cluster #3 suggests a high correlation of home dwell time and income, as the distribution patterns between CBGs in Cluster #3 and CBGs of high household income (see Figure 3a) are largely similar. North Metro Atlanta, where CBGs with high percentages of work-from-home workers and high educational levels are concentrated, exhibits a strong influence due to the stay-at-home orders, evidenced by the high concentration of CBGs in Cluster #3, a cluster with significantly increased home dwell time in March and April.

**Figure 5.**
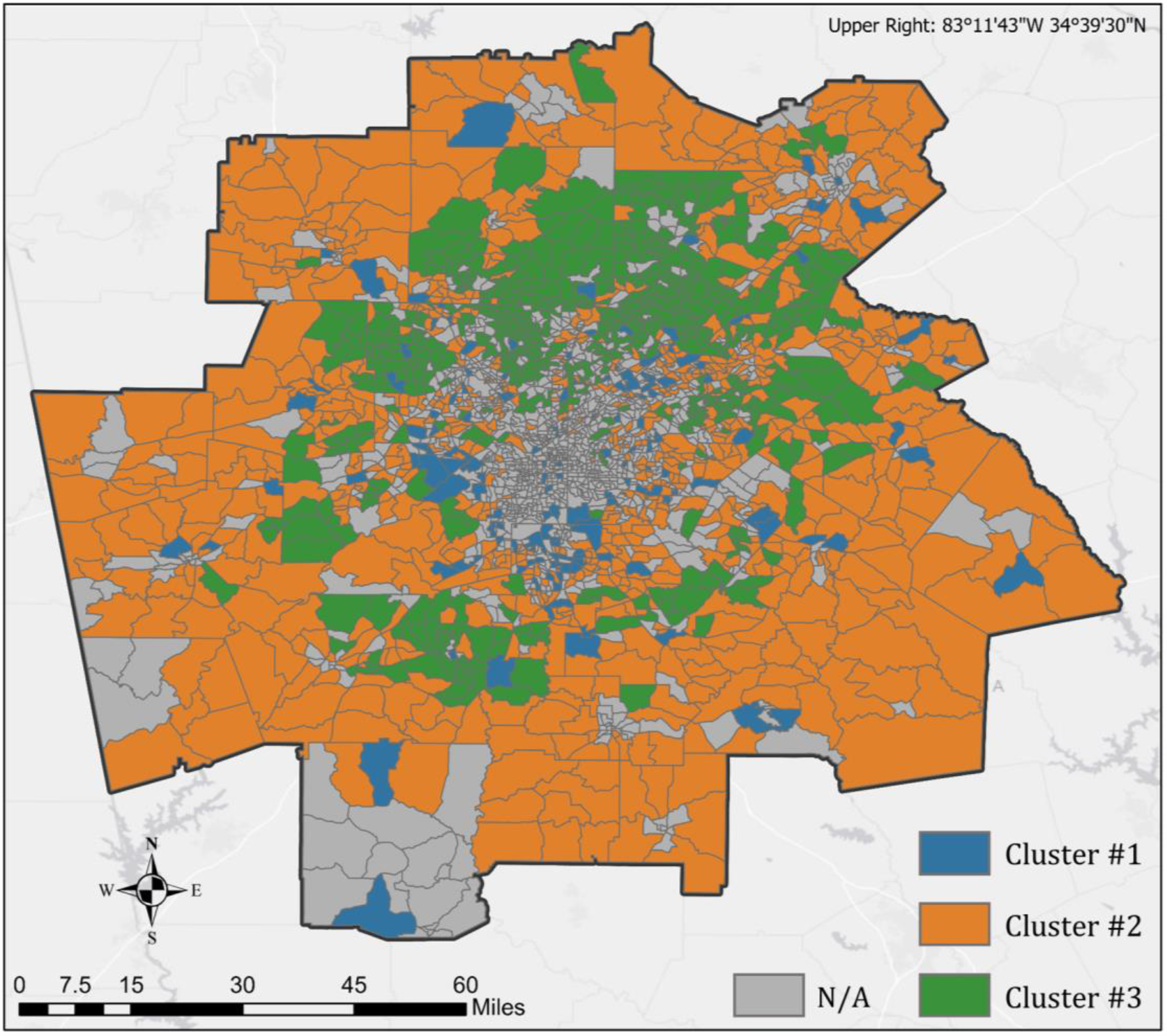
The spatial distribution of the three identified CBG clusters.

### 5.2 Demographic/socioeconomic variables in three identified clusters

The selected sixteen demographic/socioeconomic variables present unique distribution patterns in the three identified clusters (Figure 6). Compared with the other two clusters, Cluster #3 is characterized by a high median household income, a high percentage of high-earning groups, a low percentage of low-earning groups, and a low unemployment rate, suggesting that residents in rich CBGs respond to the stay-at-home order more aggressively by considerably reducing their out-of-home activities. It indicates that financial resources can, to a certain degree, influence the effectiveness of policies, as stated in other studies [16, 39].

**Figure 6.**
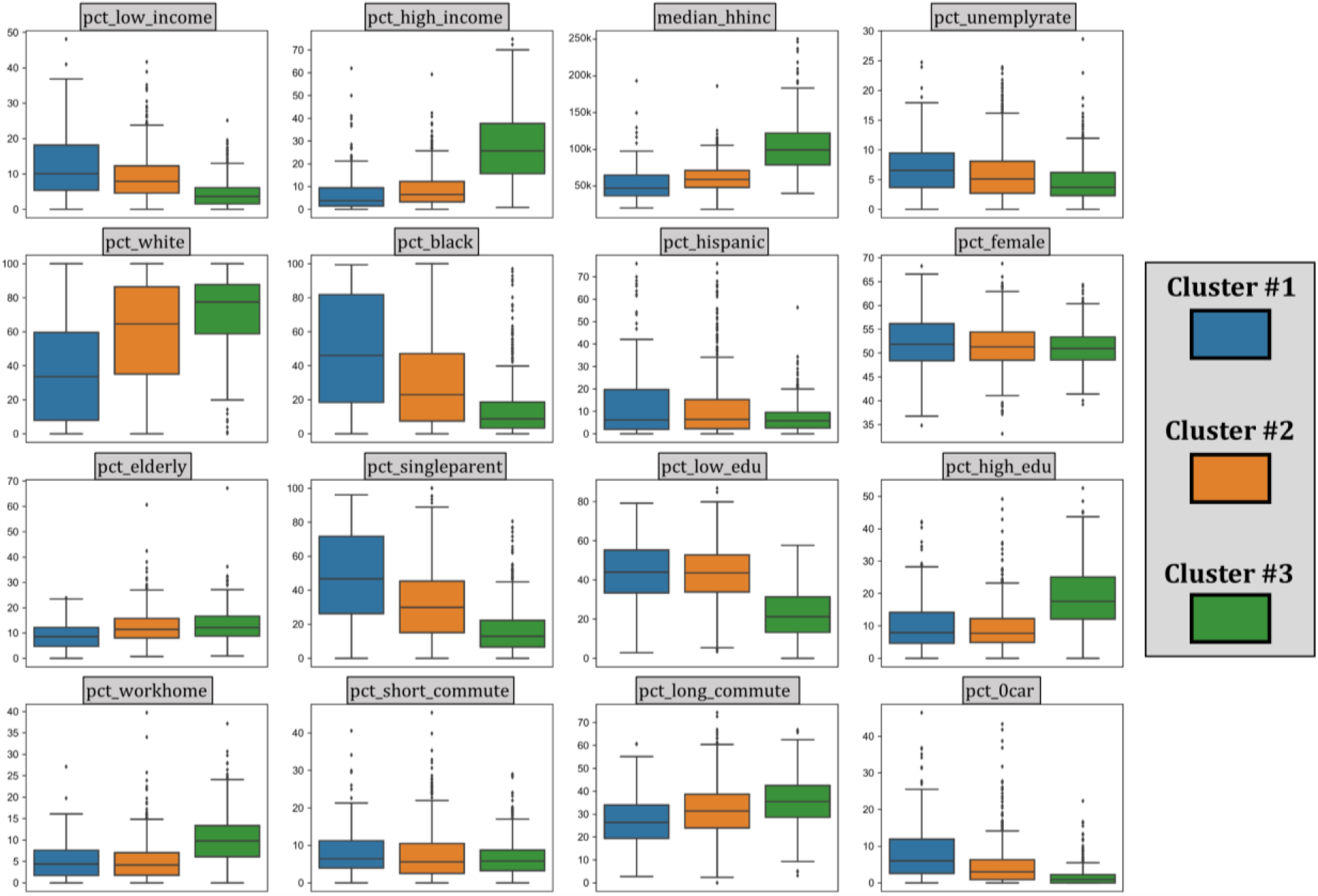
Selected demographic/socioeconomic variables in three identified clusters. The descriptions of these variables can be found in Table 1.

In terms of racial composition, the three clusters are distinctly different. The mean Black percentages of CBGs in Cluster #1, #2, and #3 are respectively 49.5%, 31.3%, and 14.5%. CBGs in Cluster #1 (with unnoticeable home dwell time increase) present much higher Black percentages than Cluster #3 (with strong home dwell time increase), revealing that stay-at-home order is less effective for CBGs with higher Black percentages. This finding coincides with other recent studies that identified the racial disparities during the COVID-19 pandemic [40, 41]. As expected, Cluster #1 also presents a higher single-parent family percentage, given the fact that a high percentage of single-parent families is usually seen in Black communities [42]. In contrast, the three identified clusters present similar Hispanic and female percentages, indicating their weaker role in distinguishing the patterns of home dwell time.

As for education, CBGs in Cluster #1 and #2 show similar distribution of the percentages of low education (42.1% and 43.1% as mean) while CBGs in Cluster #3 shows a considerably lower percentage (22.7% as mean). A reversed pattern can be found for high education, where Cluster #3 presents a notably higher percentage of high education compared to Cluster #1 and #2.

The percentages of short-commuters remain similar in all three clusters, while the percentages of long-commuters differ. The mean percentages of long-commuters in Cluster #1, #2, and #3 are 27.3%, 31.8%, and 35.4%, respectively. The result points out that a stronger increase in home dwell time is in tandem with a higher percentage of long-commuters.

### 5.3 ANOVA and Tukey’s test for clustered CBGs

We perform AVNOA to assess the statistical difference of demographic/socioeconomic variables among the three identified clusters and post-hoc Tukey’s test to evaluate the statistical difference between a certain cluster pair. The results from ANOVA suggest that all selected variables, except for the percentage of females (pct_female) and the percentage of short- commuters (pct_short_commute), show a statistically significant difference (α = 0.001) among the three clusters (Table 2). The results reveal that gender and the percentage of short-commuters are not significantly different (α = 0.001) among the means of the three identified clusters, indicating that these two variables play a weaker role in explaining the disparity in patterns of home dwell time.

**Table 2.** ANOVA results.

| Variable notations | F statistics | Sig. (p-value) |
| --- | --- | --- |
| **Economic status** |  |  |
| pct_low_income | 151.71 | <0.001* |
| pct_high_income | 516.20 | <0.001* |
| median_hhinc | 507.33 | <0.001* |
| pct_unemployrate | 32.94 | <0.001* |
| **Races and ethnicities** |  |  |
| pct_white | 108.15 | <0.001* |
| pct_black | 137.81 | <0.001* |
| pct_hispanic | 30.34 | <0.001* |
| **Gender, age, and household type** |  |  |
| pct_female | 4.98 | 0.017 |
| pct_elderly | 22.40 | <0.001* |
| pct_signleparent | 177.58 | <0.001* |
| **Education** |  |  |
| pct_low_edu | 366.83 | <0.001* |
| pct_grad_edu | 261.75 | <0.001* |
| **Transportation** |  |  |
| pct_workhome | 193.02 | <0.001* |
| pct_short_commute | 4.56 | 0.022 |
| pct_long_commute | 36.19 | <0.001* |
| pct_0car | 131.14 | <0.001* |

\* $p < 0.001$

To provide deeper insights into the comparisons of selected variables between a specific pair of clusters, we further conduct post-hoc Tukey’s test (Figure 7). For variables regarding economic status, Cluster #3 is statistically different (α = 0.001) from Cluster #1 and #2 in all four economic-related variables, i.e., pct_low_income, pct_high_income, median_hhinc, and pct_unemployrate. Cluster #1 and Cluster #2 present a weaker difference (α = 0.01) in median_hhinc and are not significantly different in pct_high_income. Results of racial and ethnic variables suggest that three clusters are statistically different from each other in pct_white, pct_black, and pct_hispanic, despite the weaker difference in pct_hispanic (α = 0.05) between Cluster #1 and #2. The difference in education (pct_low_edu and pct_high_edu) is not significant between Cluster #1 and Cluster #2 but is significant (α = 0.001) when comparing Cluster #3 to either Cluster #1 or #2. It suggests that CBGs in Cluster #3, a cluster with a strong increase in home dwell time, are characterized by their residents with high education, which is statistically different from the other two clusters. In addition, the three clusters are statistically different (α = 0.001) from each other in terms of long- commuters (pct_long_commute) and car ownership (pct_0car), suggesting that these two variables partially explain the disparity in home dwell time.

**Figure 7.**
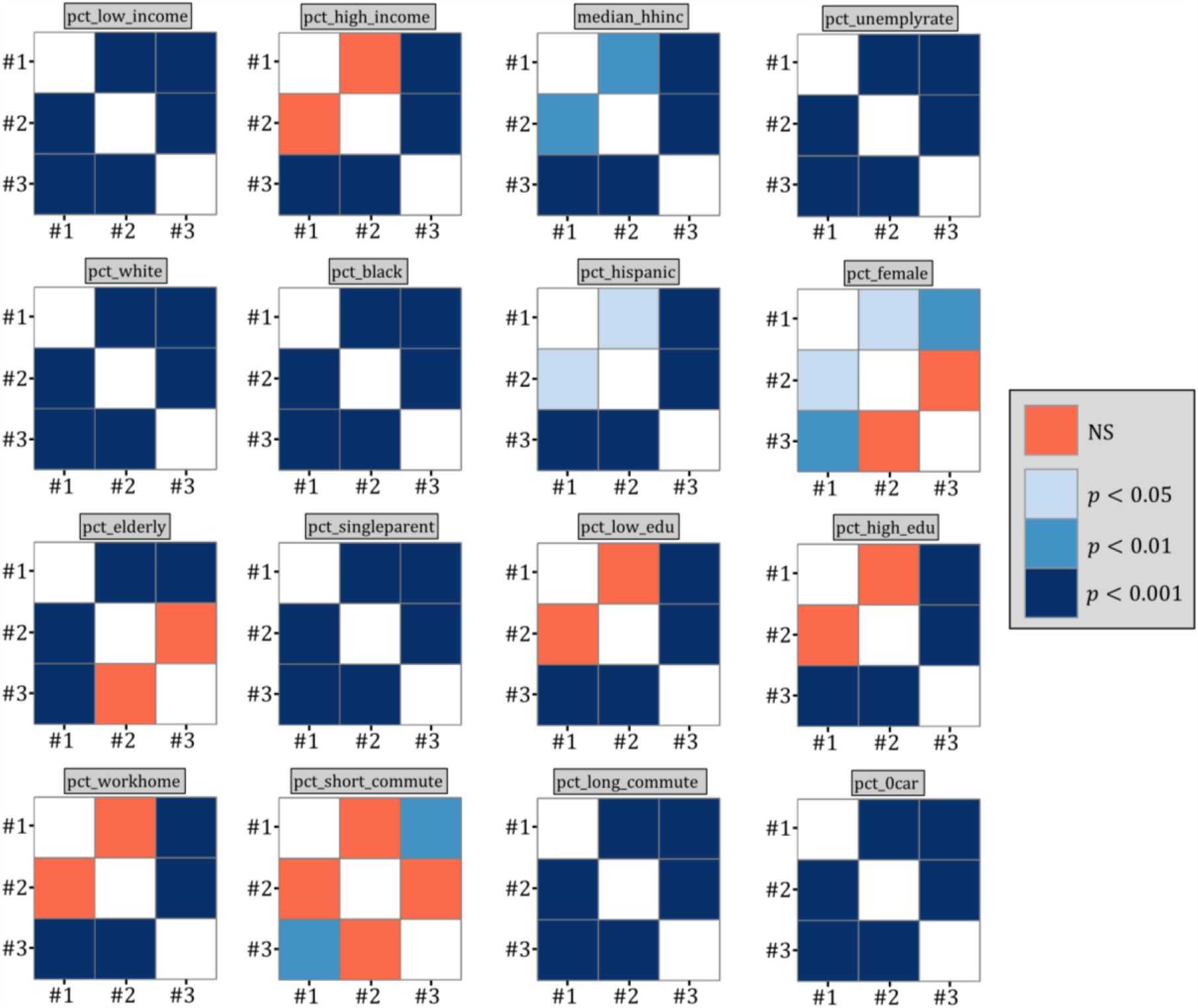
Post-hoc Tukey’s test for selected variables between pairs of clusters. NS demotes “not significant” (*p* ≥ 0.05).

## 6. Discussion

### 6.1 What do we learn?

This study applies a time-series clustering technique to categorize fine-grained mobility records (at CBG level) during the COVID-19 pandemic. Through the investigation of the demographic/socioeconomic variables in identified time-series clusters, we find that they are able to explain the disparity in home dwell time in response to the stay-at-home order, which potentially leads to disproportionate exposures to the risk from the COVID-19. This study also reveals that socially disadvantaged groups are less likely to follow the order to stay at home, pointing out the extensive gaps in the effectiveness of social distancing measures exist between socially disadvantaged groups and others. To make things worse, the existing socioeconomic status induced disparities are often exaggerated by the shortcomings of U.S. protection measures (e.g., health insurance, minimum incomes, unemployment benefits), potentially causing long-term negative outcomes for the socially disadvantaged populations [10]. In addition to the many pieces of epidemiological evidence that prove a strong relationship between social inequality and health outcomes [43, 44], this study offers evidence in the COVID-19 pandemic we are facing.

Specifically, we find that all selected variables, except for the percentage of females (pct_female) and the percentage of short-commuters (pct_short_commute), show a statistically significant difference (α = 0.001) among the three identified clusters. CBGs in Cluster #3, a cluster with strong response in home dwell time, are characterized by high median household income, high Black percentage, high percentage of high-earning groups, low unemployment rate, high education, low percentage of single parents, high car ownership, and high percentage of long- commuters. The statistically significant difference of demographic/socioeconomic variables in Cluster #3 collectively points out the privilege of the advantaged groups, usually the White and the affluent.

The weak response from the socially disadvantaged groups in home dwell time can be possibly explained by the fact that policies can sometimes unintentionally create discrimination among groups with different socioeconomic status [16], as people can react to policies based on the financial resources they have [45], which in return, influences the effectiveness of the policies. Our study reveals that the long-standing inequity issue in the U.S. stands in the way of the effective implementation of social distancing measures. Thus, policymakers need to carefully evaluate the inevitable trade-off among different groups and make sure the outcomes of their policies reflect not only the preferences of the advantaged but also the interests of the disadvantaged.

### 6.2 Limitations and future directions

It is important to mention several limitations of this study and provide guidelines for future directions. First, we acknowledge the subjectivity of predefining the number of clusters in the Kmeans clustering algorithm. In this study, we set the number of clusters as three (i.e., *k* = 3) via the investigation and interpretation of the home dwell time records from SafeGraph. We notice that, even after the preprocessing, some CBGs still present unstable temporal patterns due to the low and varying daily device count. Our interpretation of the data records reveals three distinct temporal patterns with a strong, moderate, and unnoticeable increase in home dwell time during March and April (hence, *k* is predefined as 3). To ensure the interpretability of clusters, the selection of the number of clusters in Kmeans via prior knowledge (priori) is common. However, we acknowledge that approaches like Elbow Curve [46] and Silhouette analysis [47] are largely adopted to facilitate the optimization of *k* without prior knowledge. When conducting a cross-city comparison or reproducing our approach in another region, we advise re-investigating the pattern of the time-series or adopting the aforementioned approaches to derive a reasonable setting of *k*.

Second, we construct and cluster the time-series of home dwell time using the data in the year 2020 (January 1 to Aug 31), without considering the changes in time-series compared to the previous year. It is reasonable to assume that deriving a cross-year change index facilitates the identification of CBGs that behave differently compared to the year 2019. However, we need to acknowledge the involvement of data records in the year 2019 inevitably introduces a certain level of uncertainty, as daily device count may vary substantially, leading to different representativeness of the same CBG between the two years. In addition, the Kmeans time-series clustering algorithm in this study takes the 8-month period as input. Further efforts can be directed towards the exploration of how CBGs behave differently at a certain time frame window, e.g., March and April, when strict social distancing measures were implemented.

Third, this study selects a total of sixteen variables from five major categories and explores the distribution of these variables in three identified clusters. Although previous studies have demonstrated the strong linkage between these variables and the participation of out-of-home activities, we can not rule out the possible contribution of other demographic/socioeconomic variables that are not included in this study. Future studies need to incorporate more variables to understand their roles in how social distancing guidelines are practiced. In addition, it is reasonable to assume that these variables drive the disparity in home dwell time, not independently but collectively. Therefore, statistical approaches like multinomial logit regression [48] can be used to further investigate the interactions among these variables towards time-series-based cluster generation.

Finally, it should be noted that the demographic structure, spatial pattern, and built environment vary substantially across areas, especially across densely populated urban fabrics [49,50]. Thus, the influence of demographic/socioeconomic variables on the disparity in home dwell time following the stay-at-home order may not hold the same and tend to vary geographically. In addition, local governments had differing responses to the pandemic with varying strictness of the implemented social distancing measures, potentially leading to an unequal impact that disfavors disadvantaged groups. This study only explores the situation in Metro Atlanta, which can not be generalized to other regions without caution. Thus, it is necessary to conduct comparative studies that include multiple regions to better understand the contribution of demographic/socioeconomic variables to the impact of the COVID-19 pandemic on mobility-related behaviors.

## 7. Conclusion

This study categorizes the time-series of home dwell time records during the COVID-19 pandemic, and further explores what demographic/socioeconomic variables differ among the categories with statistical significance. Taking the Atlanta-Sandy Springs-Roswell metropolitan statistical area (Metro Atlanta) as a study case, we investigates the potential driving factors that lead to the disparity in the time-series of home dwell time, providing fundamental knowledge that benefits policy-making for better mitigation measures of future pandemics.

We find that demographic/socioeconomic variables can explain the disparity in home dwell time in response to the stay-at-home order, which potentially leads to disproportionate exposures to the risk from the COVID-19. The results further suggest that socially disadvantaged groups are less likely to follow the order to stay at home, pointing out the extensive gaps in the effectiveness of social distancing measures exist between socially disadvantaged groups and others. Specifically, we find that CBGs with strong response to the stay-at-home order are characterized by high median household income, high Black percentage, high percentage of high-earning groups, low unemployment rate, high education, low percentage of single parents, high car ownership, and high percentage of long-commuters, pointing out the privilege of the advantaged groups, usually the White and the affluent. In other words, populations with lower socioeconomic status may lack the freedom or flexibility to stay at home, leading to the exposure of more risks during the pandemic. Our study reveals that the long-standing inequity issue in the U.S. stands in the way of the effective implementation of social distancing measures. Thus, policymakers need to carefully evaluate the inevitable trade-off among different groups and make sure the outcomes of their policies reflect not only the preferences of the advantaged but also the interests of the disadvantaged.

## Data Availability

The original data is derived from SafeGraph. The preprocessed data can be obtained upon request.

## Author Contributions

Conceptualization, Xiao Huang, Sicheng Wang, Hanxue Wei, and Baixu Cheng; Methodology, Xiao Huang, Junyu Lu, Sicheng Wang, Hanxui Wei, and Baixu Chen; Formal Analysis, Xiao Huang, Junyu Lu; Writing-Original Draft Preparation, Xiao Huang, Sicheng Wang, Hanxue Wei; Writing-Review & Editing, Xiao Huang, Junyu Lu, Zhenlong Li; Supervision, Xiao Huang, Zhenlong Li; Funding Acquisition, Xiao Huang.

## Acknowledgments

The authors want to thank SafeGraph for providing the home dwell time dataset, which makes this research possible.

## Appendix A

**Table A.** U.S. counties that are included in Metro Atlanta.

| **GEOID<sup>1</sup>** | **County name** |
| --- | --- |
| 13171 | Lamar County |
| 13063 | Clayton County |
| 13089 | DeKalb County |
| 13227 | Pickens County |
| 13045 | Carroll County |
| 13297 | Walton County |
| 13013 | Barrow County |
| 13223 | Paulding County |
| 13199 | Meriwether County |
| 13113 | Fayette County |
| 13143 | Haralson County |
| 13015 | Bartow County |
| 13139 | Hall County |
| 13077 | Coweta County |
| 13159 | Jasper County |
| 13151 | Henry County |
| 13085 | Dawson County |
| 13097 | Douglas County |
| 13211 | Morgan County |
| 13135 | Gwinnett County |
| 13231 | Pike County |
| 13247 | Rockdale County |
| 13121 | Fulton County |
| 13149 | Heard County |
| 13067 | Cobb County |
| 13057 | Cherokee County |
| 13217 | Newton County |
| 13255 | Spalding County |
| 13035 | Butts County |
| 13117 | Forsyth County |

<sup>1</sup>GEOID is a 5-digit county code defined by the U.S. Census Bureau.

